# Impact of reference electrode position on motor unit number estimation (MUNE) in the tibialis anterior muscle using MScanFit: test-retest reliability

**DOI:** 10.1101/2023.09.20.23295858

**Authors:** M Almokdad, BG Yang, B Jantz, A Abrahao, KE Jones

**Affiliations:** Faculty of Kinesiology, Sport, & Recreation, University of Alberta, Edmonton, Canada; Department of Medicine, University of Toronto, Toronto, Canada; Neuroscience and Mental Health Institute, University of Alberta, Edmonton, Canada

**Keywords:** electromyography, coefficient of repeatability, humans, electrical stimulation, skeletal muscle, compound muscle action potential, lower motor neuron

## Abstract

**Objectives:** This study aimed to assess the effect of varying the reference electrode position, specifically comparing position A3 (medial patella) to routine position 1 (R1) and the MScan multicenter protocol position (M1), on compound muscle action potential (CMAP) and motor unit number estimation (MUNE) in the tibialis anterior muscle of healthy participants.

**Methods:** Twenty healthy participants underwent repeated MScanFit MUNE assessments with a 7-14 day interval between tests. The reference electrode (E2) was placed in three positions at each visit (A3, R1, and M1), while the active electrode (E1) remained constant. An additional seventeen participants were included to establish the minimal detectable true change in MUNE values using MScanFit, with the reference electrode exclusively in the M1 position.

**Results:** The reference electrode position significantly influenced CMAP and MUNE, with R1 resulting in lower values. However, no significant difference was observed between M1 and A3 positions. Relative and absolute reliability indicators favored using the M1 position for reference in MScanFit MUNE. In a combined dataset of 37 healthy participants, the average tibialis anterior muscle motor unit count was estimated at 148 (SD 25.2), with a minimal detectable true change of 55 units.

**Conclusions:** The preference for the M1 position over the alternative A3 position is supported, particularly for MScanFit MUNE assessments in the tibialis anterior muscle. Clinically, a true change in MUNE should consider the minimal detectable change of 55 motor units, underscoring the reality that large changes in MUNE are required to conclude a genuine change beyond measurement error.

**Significance:** For MUNE examinations of the tibialis anterior muscle, adhering to the electrode positions outlined in the MScan multicenter protocol is advisable. Awareness of measurement error limitations in MScanFit MUNE underscores its applicability in making longitudinal clinical decisions for *individual* patients.

## 1. Introduction

Motor unit number estimation (MUNE) is a technique combining electromyography and peripheral nerve stimulation to quantify the motor unit count within skeletal muscles. The method was first introduced by McComas and colleagues in 1971, and subsequent years have seen the development of various alternative approaches (as comprehensively reviewed by Gooch et al., 2014; de Carvalho et al., 2018; Wright et al., 2023). Among these approaches is MScanFit MUNE, proposed by Bostock in 2016, which shares many similarities with standard nerve conduction studies and is currently undergoing evaluation in international multicenter studies to assess its measurement properties as a lower motor neuron biomarker for conditions like amyotrophic lateral sclerosis (ALS; Sørensen et al., 2022; Sørensen et al., 2023).

A recent study involving 15 participating centers from nine countries reported discrepancies in standard measures of compound muscle action potential (CMAP) amplitudes, despite all centers adhering to the same standard operating procedure (Sørensen et al., 2023). Our current study seeks to investigate whether these discrepancies could have been mitigated by utilizing an alternative (optimal) reference electrode position when examining the tibialis anterior muscle. The tibialis anterior muscle was chosen because it has received particular attention with regard to electrode position for motor nerve conduction studies (Escorcio-Bezerra et al., 2019; Nandedkar and Barkhaus, 2020).

The amplitude of the CMAP hinges on the precise placement of three surface electrodes: the active (E1), the reference (E2), and the differential amplifier ground (E0, Dumitru et al., 2002). Nerve conduction studies, including the MScan Multicenter protocol, typically employ a monopolar or belly-tendon electrode configuration. In this setup, the E1 electrode is positioned over the muscle belly and adjusted to optimize the negative peak of the CMAP—a position operationally defined as the motor point (Bowden and McNulty, 2012). The loquacious E0 electrode (Robinson et al., 2016) is situated on the ipsilateral limb, usually over a bony landmark. In the MScan Multicenter protocol, the E2 electrode is placed over the distal tendon. Notably, electrode placement significantly influences the size and shape of CMAPs in nerve conduction studies (Dumitru and King, 1991; van Dijk et al., 1994; Falck and Stalberg, 1995; Nandedkar and Barkhaus, 2020). In the context of motor nerve conduction to the tibialis anterior muscle via the fibular nerve, the optimal position for the E2 electrode has been identified as the medial aspect of the ipsilateral patella (Escorcio-Bezerra et al., 2019; Nandedkar and Barkhaus, 2020).

Our hypothesis is twofold: first, we anticipate that when conducting an MScanFit MUNE examination on the tibialis anterior muscle, the amplitude of the CMAP will be maximized and demonstrate improved reliability when utilizing the alternative E2 position on the patella compared to the position stipulated by the MScan Multicenter protocol. Second, we hypothesize that the alternative E2 position will enhance the reliability indices of motor unit number estimates in this particular muscle.

## 2. Methods

Two cohorts of healthy control participants were recruited for this study through convenience sampling. The inclusion and exclusion criteria were adopted from the larger MScan Multicenter protocol (Sørensen et al., 2023). Potential participants underwent a pre-screening process to identify and exclude individuals meeting any of the following exclusion criteria: a history of prior injuries or diseases affecting the central nervous system, leg injuries resulting in numbness or weakness, previous chemotherapy treatments, a diagnosis of diabetes mellitus, self-reported alcohol consumption exceeding 14 units per week, other causes of polyneuropathy, documented history of nerve entrapment syndromes, cognitive impairment, or pregnancy. All participants completed the questionnaire component of the Michigan Neuropathy Screening Instrument, followed by a neurological examination (Feldman et al., 1994).

The study involved two cohorts:

1. The first cohort consisted of 17 individuals (self-reported sex: 6 male and 11 female), recruited during the summer of 2021, within an age range of 18-80 years. Notably, nine of these participants had previously participated in a related study (Sorensen et al., 2023). All electrodiagnostic testing in this cohort was conducted by the same person (Rater #1) under the mentoring and supervision of a clinical neurophysiologist.
2. The second cohort comprised 20 individuals (14 male and 6 female), recruited during the summer of 2023, with an age range of 18-40 years. Electrodiagnostic testing for this cohort was carried out by a second person (Rater #2) after receiving training from a clinical neurophysiologist.

Each participant attended two laboratory visits (visit 1 & 2), with these visits separated by a 7-14 day interval. Informed written consent was obtained from all participants, and the study was approved by the Research Ethics Board of the University of Alberta (Protocol#: Pro00061945), conducted in adherence to the Declaration of Helsinki.

### 2.1 Surface Electrode Placements

Participants were positioned in a semi-reclined posture on an examination chair, with their feet elevated. To maintain a consistent ankle angle of approximately 90 degrees, an ankle-foot orthosis (AFO) was used on the tested limb, complemented by a pillow placed behind the knees. (Note: CMAP amplitude is dependent on ankle angle and consistent positioning using the AFO was expected to improve repeatability (Frigon et al., 2007).) The skin was meticulously marked, and measurements were taken to standardize electrode placement.

Specifically, the head of the fibula was palpated and marked, followed by the measurement of the distance from the head of the fibula to the medial malleolus. A mark was made on the skin at one-third of this measured distance distal to the head of the fibula, designating it as the starting position for the active (E1) electrode.

The reference electrode (E2) was situated in one of three positions following the MScan Multicenter protocol, along with routine and alternative positions as used by Escorcio-Bezerra et al. (2019):

- **M1**: The E2 electrode was placed at the position specified in the MScan Multicenter protocol, marked on the skin 5 cm proximal to the intermalleolar line along the tendon of the tibialis anterior muscle.
- **R1**: A routine position utilized in motor nerve conduction studies was determined by measuring the length of the tibia (from the tibial plateau to the intermalleolar line) and marking the skin over the tibia at two-thirds of this distance from the tibial plateau.
- **A3**: The alternative, optimal position for the reference electrode was located at the midpoint on the medial aspect of the ipsilateral patella. (Note: this same position was given the label P by Nandedkar and Barkhaus, 2020.)

The E0 (differential ground) electrode was placed over the tibia, situated between the head of the fibula and the initial mark designated for the E1 electrode. The position for the stimulating cathode was determined through percutaneous stimulation using a handheld probe (9 mm diameter) just below the head of the fibula. The selected position was one with the lowest electrical threshold, eliciting a palpable twitch in the tibialis anterior muscle when subjected to a 0.5 ms pulse. The return anode for stimulation was situated approximately 5-6 cm proximal to this selected site, near the lateral femoral condyle. A visual representation of this placement can be found in a prior publication (Fig 1C, Sørensen et al., 2022).

To prepare the skin at each electrode site, a process of hair removal, gentle abrasion (using gel or fine-grain sandpaper), and alcohol swab cleaning was meticulously followed. Impedance between any two electrodes was monitored to achieve a skin-electrode impedance falling within the range of 5 - 10 kOhms. The electrodes employed were consistent with those specified in the MScan Multicenter protocol, including stimulation electrodes ([Ambu® BlueSensor QR](https://www.ambu.com/cardiology/ecg-electrodes/product/ambu-bluesensor-qr)), the E1 and E2 recording electrodes ([Ambu® BlueSensor NF](https://www.ambu.com/cardiology/ecg-electrodes/product/ambu-bluesensor-nf), dimensions), and the E0 electrode ([Ambu® Neuroline™ Ground](https://www.ambu.com/neurology/emg-electrodes/product/ambu-neuroline-ground)). Skin temperature was carefully maintained between 32 - 34 degrees Celsius, with the utilization of heat packs when necessary.

### 2.2 Recording and MScanFit Procedure

The recording electrodes interfaced with a D440 amplifier ([Digitimer Ltd.](https://www.digitimer.com/product/life-science-research/amplifiers/d440-2-or-4-channel-isolated-amplifier/), gain 300-500, bandpass filter settings 5 Hz - 10 kHz). The amplifier’s output was subsequently passed through a 60 Hz HumBug noise eliminator ([Digitimer Ltd.](https://www.digitimer.com/product/life-science-research/mains-noise-eliminators/humbug-noise-eliminator/)) before being routed to an isolated, multifunction DAQ device (National Instruments USB BNC-6251). Data acquisition was conducted at a sampling frequency of 10 kHz using QtracW software (©University College London, U.K. distributed by [Digitimer Ltd](https://www.digitimer.com/product/life-science-research/software/qtracw-threshold-tracking-software/).). The stimulating electrodes were connected to a DS5 bipolar constant current stimulator ([Digitimer Ltd.](https://www.digitimer.com/product/human-neurophysiology/peripheral-stimulators/ds5-isolated-bipolar-constant-current-stimulator/)) which was controlled via analogue inputs from the DAQ device, all under the control of the QtracW software.

The automation of the MScanFit MUNE examination was facilitated by employing the MScan-R2 Qtrac procedure file, a method detailed in prior works (Sorensen et al., 2022; Sorensen et al., 2023; the Qtrac procedure file used has been deposited to Figshare https://doi.org/10.6084/m9.figshare.24112533.v1). The stimulus duration was 0.5 ms, as per the MScan Multicenter protocol for testing the common fibular nerve to the tibialis anterior muscle.

The stimulus amplitude was manually adjusted by the operator to determine the level necessary for generating the maximum CMAP. Following this initial determination, the E1 electrode was methodically repositioned to explore the configuration that produced the highest CMAP amplitude (measured from baseline to negative peak). The E1 electrode underwent at least three adjustments, involving both proximal-distal and medial-lateral movements, to pinpoint the optimal position. This position remained consistent throughout the data collection process.

In cohort #1, the E2 electrode was placed in the standard MScan Multicenter position, referred to as M1. Conversely, in cohort #2, participants had the E2 electrode positioned in one of three locations: M1, R1, or A3. To introduce randomness into the sequence of testing, twenty permutations of these three positions were pre-generated and used for both the first visit and the subsequent visit scheduled seven to fourteen days later.

The procedure for recording the comprehensive stimulus-response, or CMAP scan, entailed a rapid increase in stimulus amplitude to achieve a supramaximal level. In cohort #2 data collection, the same supramaximal stimulus level was uniformly applied for all three E2 positions during a study visit. Subsequent to establishing the supramaximal level, a series of 20 pre-scan stimuli were administered at this amplitude, with interstimulus intervals maintained between 0.6 to 1.0 seconds. Subsequently, each successive stimulus amplitude was decreased by 0.2% until it was deemed subthreshold by the operator. To gauge instrument variance in the absence of an evoked response, a final set of 20 post-scan stimuli was delivered at the subthreshold intensity.

For participants in cohort #2, this entire process was repeated three times, with the E2 electrode repositioned to all three designated positions. Participants returned for a retest study visit seven to fourteen days later to determine reliability.

### 2.3 MScanFit MUNE Analysis

For the analysis of the CMAP scan data, QtracW software was employed, following established procedures (Bostock, 2016; Sørensen et al., 2022). The initial step in the analysis involved manual delineation of the pre-scan and post-scan limits. This process aimed to identify a range of CMAP amplitudes representing the maximal plateau value (pre-scan) and a range of subthreshold responses devoid of potential confounding factors such as voluntary contractions or inadvertent evoked responses (post-scan).

Subsequent to this step, a preliminary analytical model was constructed based on slope and variance observed during the CMAP scan (Bostock, 2016). The final phase of the MScanFit analysis was an iterative optimization process, where the model was continuously refined to align with the experimental data. QtracW software provides flexibility in parameter selection and optimization methods. The following settings were utilized:

- Number of repeats of modeled scans: Auto
- Amplitude resolution: 40 uV
- Max number of units: 300
- Maximum time for fitting (minutes): 10
- Minimum size for units: Absolute, 5 uV, %CMAP amp, 0.333, %SD noise, No minimum, %mean unit amp, 20
- Optimization option: All Best of n
- RS (Rejection Sampling): 2.0
- Optimization method: Method 2

To estimate the variance in the final MUNE outcome measure generated by MScanFit, the model fit and optimization process were repeated twelve times for each study visit in each cohort. For instance, in cohort #1, comprising 17 participants with two visits each, this resulted in 408 repeated MScanFit model fits.

MScanFit analysis generated multiple outcome variables, but for our *a priori* hypotheses, we focused on examining only the peak CMAP and the MUNE value from each visit. The anonymized individual participant data has been deposited to Figshare (https://doi.org/10.6084/m9.figshare.24112533.v1).

### 2.4 Statistical Analysis

Our *a priori* alternative hypotheses centered on the influence of the reference electrode position (E2) on both CMAP amplitude and MUNE values, as well as its impact on test-retest reliability. To be more specific, we hypothesized that the proximal position for the E2 electrode (A3) would produce the largest CMAP amplitudes and most reliable metrics for MUNE. Based on the empirical correlation between CMAP and MScanFit MUNE (e.g. Pearson’s r = 0.80 for the tibialis anterior muscle Sørensen et al., 2022), the expectation was that MUNE values would also be highest when using the A3 position for the reference electrode.

The distribution of the two dependent variables (CMAP and MUNE) were assessed for normality with a Shapiro-Wilk’s test; a threshold alpha of 0.05 was used to reject the null hypothesis and prompt further evaluation. Following assessment of the distribution of the data, a two-way repeated measures ANOVA (visit: two levels, and E2 position: three levels) was run to identify any interaction and main effects. Mauchly’s test was used to test the assumption of sphericity, and the Greenhouse-Geisser correction was used when this assumption was violated. All analysis for null hypothesis significance testing was done using IBM SPSS Statistics (Version 29). Pairwise comparisons and data visualization used permutation tests and estimation statistics (Ho et al., 2019).

Both relative and absolute reliability indices were calculated to provide metrics for evaluation of MScanFit MUNE as an assessment tool for clinical decision making on individual patients over time (Vaz et al., 2013). The relative reliability index was the intraclass correlation coefficient (ICC, 2-way mixed-effects model, for absolute agreement) and we used the following thresholds for qualitative categories: <0.5 poor reliability, 0.5 to 0.75 moderate reliability, 0.75 to 0.9 good reliability, and values greater than 0.90 indicate excellent reliability (Koo & Li, 2016). The relative values of ICC facilitate comparisons across measures with different units, like CMAP (mV) and MUNE (# of units). Calculations for ICC were done in MATLAB (Release 2022b, The MathWorks, Inc., Natick, Massachusetts, United States).

The chosen absolute reliability index is known synonymously as: the Coefficient of Repeatability (CR), the Smallest Real Difference, or the Minimal Detectable True Change (Vaz et al., 2013).

The advantage of using CR is immediate interpretability of the observed change in repeat measures from the same individual. This allows setting a threshold for detecting a true change in the measurement over and above the random error associated with the assessment tool and was calculated at the 95% confidence limit as,

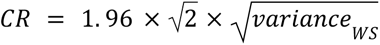

where *variance_WS_* is the average within subject variance calculated for each participant across visit 1 and visit 2. It’s worth noting that the CR calculation can be adjusted to a 90% or a 99% confidence limit by replacing the value of 1.96 with 1.645 or 2.576, respectively. The square root of *variance_WS_* is also known as the Standard Error of Measurement.

## 3. Results

A total of seventy-seven MScanFit MUNE investigations were conducted across two cohorts of participants. In cohort #1, one potential participant did not tolerate the stimulation, leading to discontinuation of the visit and subsequent data removal. Additionally in cohort #1, achieving confident supramaximal stimulation proved challenging for a second participant, likely due to the large lower leg girth and the limited current output available from the DS5 stimulator (50 mA). In cohort #2, one participant could not schedule the second visit within the 7-14 day interval, necessitating a third visit to maintain the specified timing between test and retest study visits.

### 3.1 Effect of E2 Electrode Position on CMAP and MUNE

Figure 1 presents data from a single participant during both visits, encompassing all three reference electrode positions. This participant’s data was chosen because the differences in test-retest outcome measures closely approximated the cohort #2 average, representing typical data rather than exceptional.

**Figure 1.**
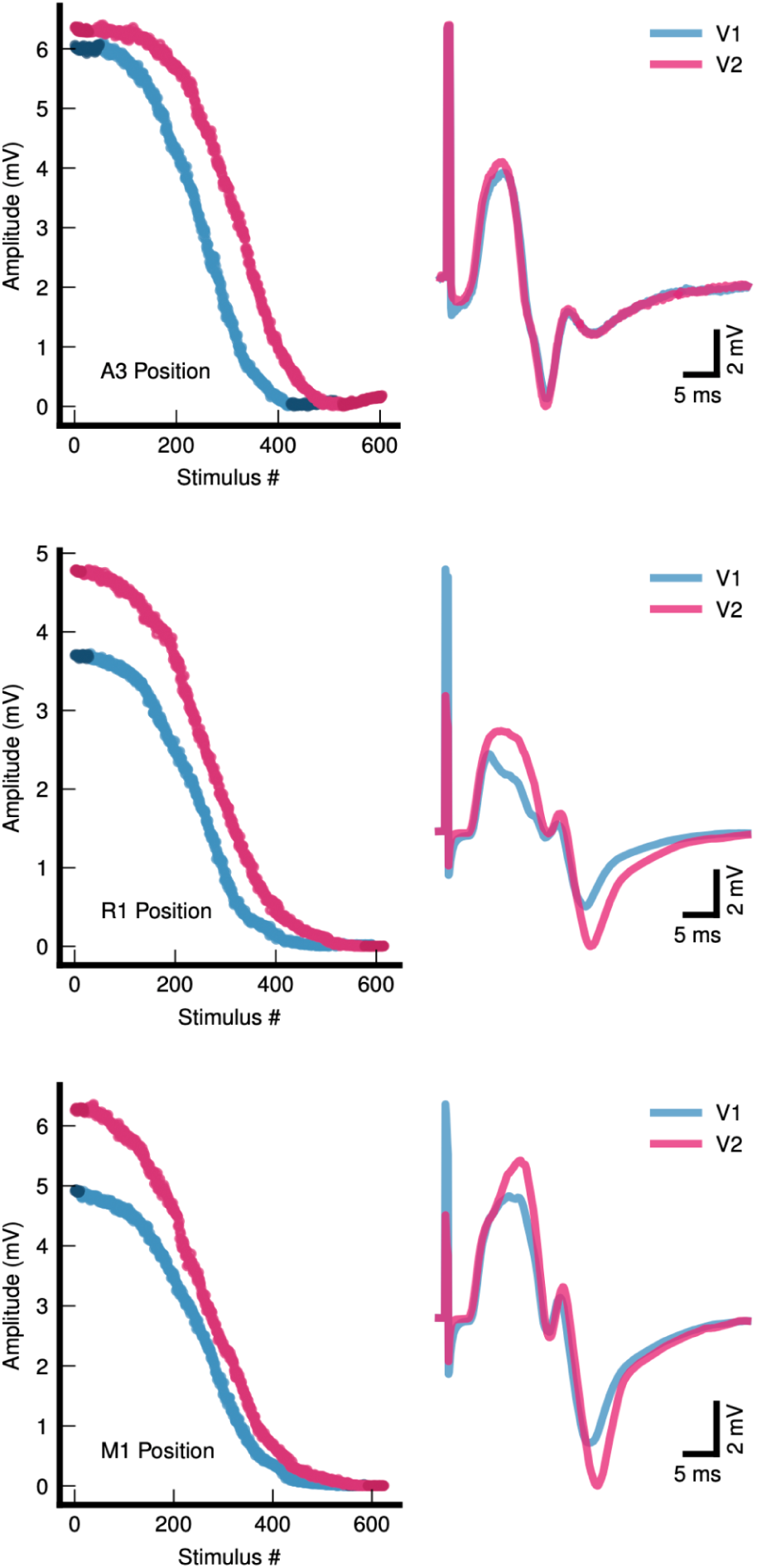
Individual data from a participant whose differences between visits was similar to the mean difference across all participants. Each row represents data for a different reference electrode position: M1, standard position for the MScan Multicenter protocol; R1, routine position used in motor nerve conduction studies; A3, alternative position demonstrated as optimal in a prior study (Escorcio-Bezerra et al., 2019). Visit 1 is shown in blue and visit 2 is shown in purple. The CMAP scans (left column) are consecutive stimulus numbers (from supramaximal to subthreshold) and CMAP size in mV. The darker colors at the start and end of the CMAP scan represent the pre-scan and post-scan sections of the data. The CMAP waveforms (right column) are shown with the stimulus artifact to illustrate an anecdotal finding about the nature of the artifact with the A3 position. The artifact did not return to baseline after the stimulus, but extended for a duration to overlap with the latency of onset of the evoked CMAP.

A two-way repeated measures ANOVA revealed no statistically significant interaction between visit and electrode position on CMAP amplitude (F(1.24,23.54) = 1.566, p = .227, using the Greenhouse-Geisser method to correct for rejection of the sphericity assumption). Furthermore, no statistically significant main effect of visit on CMAP values was detected (F(1,19) = .918, p = .350, parietal eta^2 = .046). However, a statistically significant main effect of electrode position on CMAP values was observed (F(2,38) = 18.550, p < .001, partial eta^2 = .494). Pairwise comparisons indicated that CMAP values using the R1 position were significantly smaller than those obtained with the A3 and M1 positions. Importantly, no significant difference was found in CMAP values between the A3 and M1 positions (Figure 2, left panel).

**Figure 2.**
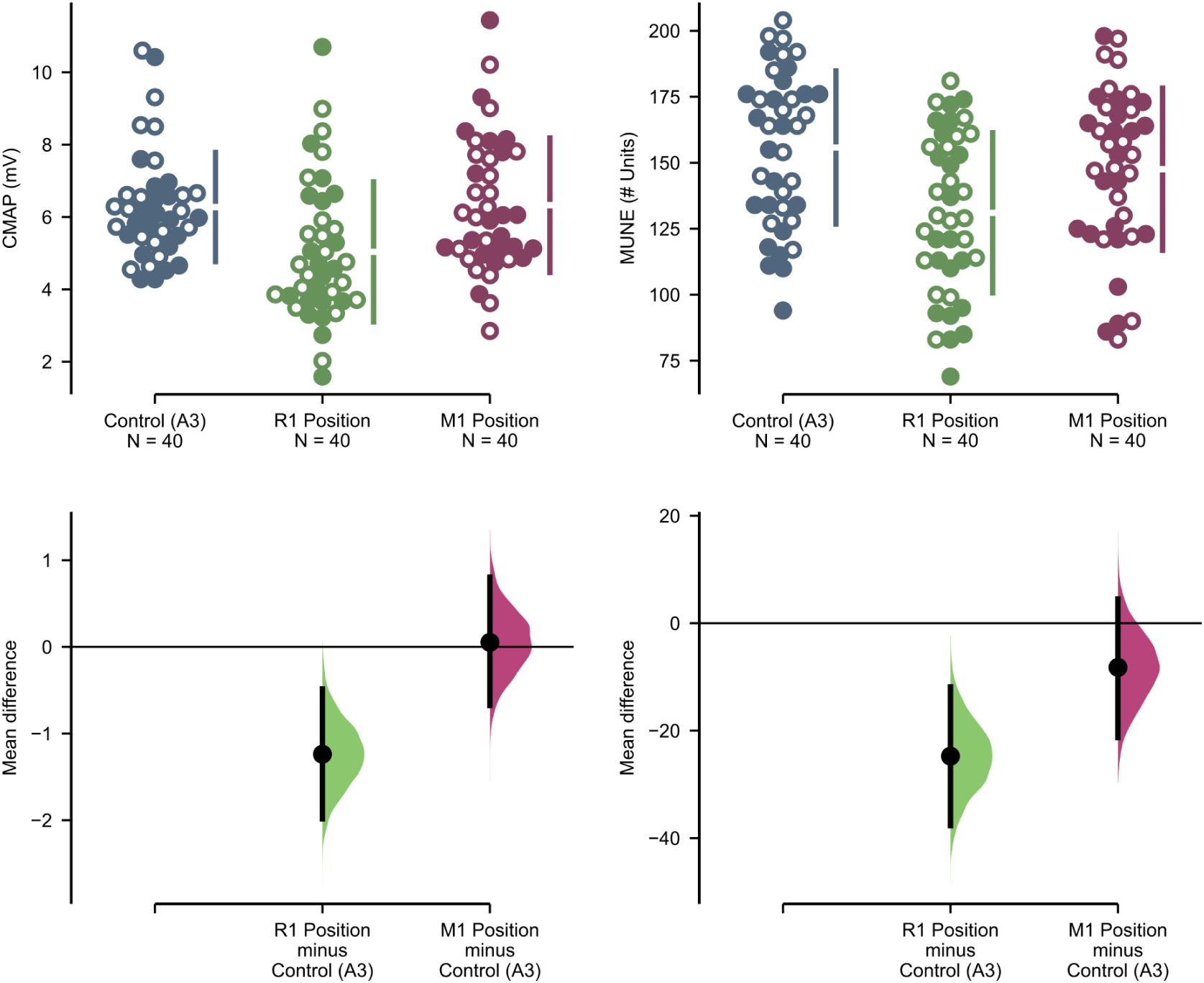
Group data from cohort #2 to show the effect of E2 electrode position on CMAP (left) and MUNE (right). Since there was no main effect of visit, both visits from the sample of 20 participants are shown together (solid symbols are V1, hollow are V2). The mean difference for 2 comparisons against the shared control Control (A3) are shown in the above Cumming estimation plot. The raw data is plotted on the upper axes. The axes on the lower row illustrate mean differences are plotted as bootstrap sampling distributions. Each mean difference is depicted as a dot with the 95% confidence interval indicated by the vertical error bars. Use of the R1 position resulted in a smaller CMAP (compared to A3: mean difference of 1.27 mV [95%CI, .51 to 1.94], p < .001; compared to M1: mean difference of 1.28 mV [95%CI, .83 to 1.73], p < .001). There was no difference in CMAP for the A3 and M1 positions (mean difference is .05 mV [-.62 to .72], p = 1.0). Similarly, the R1 position resulted in smaller MUNE values (compared to A3: mean difference is 24 units [95%CI 12 to 36], p < .001; compared to M1: mean difference is 16 units [95%CI 8. to 23], p < .001). There was no difference in MUNE when comparing the A3 and M1 positions (mean difference is -8 units [95%CI -20 to 4], p = .244).

Similarly, a two-way repeated measures ANOVA found no statistically significant interaction effect on MUNE (F(2,38) = .619, p = .544). Although there was no statistically significant main effect of visit on MUNE (F(1,19) = 3.729, p = .069, parietal eta^2 = .164), a statistically significant main effect of electrode position on MUNE was evident (F(2,38) = 18.206, p < .001, partial eta^2 = .489). Once again, pairwise comparisons showed that MUNE values were lower when using the R1 position, while values were comparable between the A3 and M1 positions (Figure 2, right panel).

### 3.2 M1 Position Favored for Reliability

Relative reliability for the CMAP was moderate across all three electrode positions (Table 1, ICC values range from 0.55 to 0.72), and absolute reliability was similar across these positions (Table 1, the average Coefficient of Repeatability (CR) was 2.9 mV). However, relative reliability for MUNE was moderate in the M1 but poor with the R1 and A3 positions (Table 1, ICC).

**Table 1.** Comparison of measures of CMAP and MUNE in the same participants using the three different reference electrode positions. All measures performed by Rater 2.

| Position | Measure | Visit 1 |  | Visit 2 |  | Relative and absolute reliability indices |  |  |  |  |  |  |  |
| --- | --- | --- | --- | --- | --- | --- | --- | --- | --- | --- | --- | --- | --- |
|  |  | Mean | SD | Mean | SD | ICC | Mean diff (bias) [95% CI] | SD<sub>diff</sub> between | p-value | 95% LOA - lower | 95% LOA - upper | Within subject variance | CR (95%) |
| M1 | CMAP (mV) | 6.41 | 1.90 | 6.24 | 1.86 | 0.67 | -0.17 [-0.95, 0.39] | 1.54 | 0.640 | -2.85 | 3.19 | 1.14 | 2.96 |
|  | MUNE | 144 | 30.7 | 151 | 31.0 | 0.57 | -7.5 [-4.9, 19.8] | 28.4 | 0.255 | -63.2 | 48.2 | 411.9 | 56.2 |
| R1 | CMAP (mV) | 4.96 | 2.11 | 5.12 | 1.79 | 0.72 | 0.16 [-0.59, 0.73] | 1.49 | 0.638 | -3.08 | 2.77 | 1.07 | 2.87 |
|  | MUNE | 126 | 33.7 | 136 | 26.6 | 0.46 | 9.6 [-2.5, 24.2] | 31.1 | 0.197 | -70.4 | 51.3 | 504.1 | 62.2 |
| A3 | CMAP (mV) | 5.97 | 1.39 | 6.58 | 1.60 | 0.55 | 0.60 [-0.03, 1.21] | 1.37 | 0.067 | -3.29 | 2.09 | 1.07 | 2.87 |
|  | MUNE | 148 | 30.0 | 163 | 26.6 | 0.25 | 15.2 [-0.4, 28.5] | 34.2 | 0.067 | -82.1 | 51.8 | 669.3 | 71.7 |

ICC Intraclass correlation coefficient: two-way mixed effect model (absolute agreement definition).
Mean diff (bias): mean difference between visit 1 and 2, also known as bias in Bland-Altman plots, with 95% confidence interval.
SD<sub>diff</sub> between: standard deviation of the between subject difference in values at visit 1 and 2.
The reported p-value was generated by a two-sided permutation t-test (using 5000 bootstrap samples) comparing paired data from visit 1 and 2.
95% LOA - lower & upper: Bland-Altman 95% confidence intervals of the Limits Of Agreement, for lower and upper boundaries.
CR: Coefficient of Repeatability, 95% confidence interval, $=1.96 \cdot \sqrt{2} \cdot \sqrt{[\text{within subject variance}]}$ , or $= 2.77 \cdot \text{Standard Error of Measurement (SEM)}$

Absolute reliability was superior with the M1 position, as indicated by a lower CR compared to the other positions (Table 1, CR).

The difference in test-retest reliability of CMAP and MUNE for the three electrode positions is illustrated in Figure 3. No significant bias was observed between measures on visit 1 compared to visit 2 for any of the measures (Table 1, Mean difference and p-value). Notably, while the boundaries for the limits of agreement were narrowest for CMAP using the A3 position, they were widest for MUNE in the same position, indicating greater variance between visit 1 and visit 2 (Figure 3).

**Figure 3.**
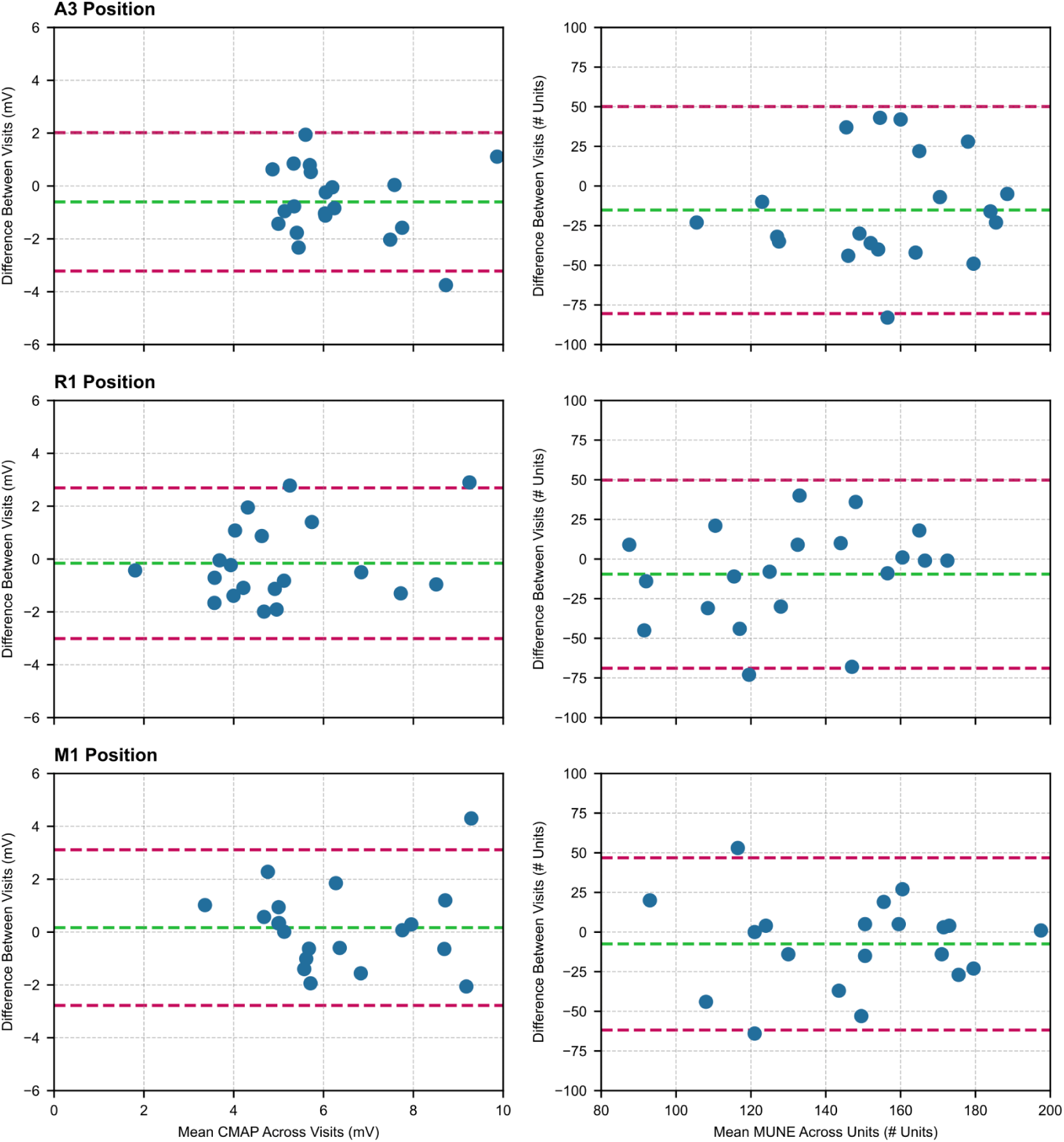
Bland and Altman difference plots for the CMAP (left) and MUNE (right) outcome measures in the three different E2 electrode positions. The horizontal and vertical axis have the same scales to allow direct visual comparison across positions. The green line represents the bias, or mean difference, while the purple lines above and below the green line represent the boundaries of the 95% confidence intervals of the limits of agreement (LOA). The numerical values for the bias and the 95% confidence intervals for the bias, and the values for the lower and upper bounds of the LOA are given in Table 1. The boundaries of the LOA for the CMAP in position A3 are narrow, however this position has the widest boundaries for MUNE.

### 3.3 MUNE Reliability Independent of Rater

Relative and absolute reliability indices for CMAP (using the M1 position) by Rater 2 were moderate to poor compared to Rater 1, suggesting potential data quality limitations (Table 2, ICC and CR for CMAP). For instance, the coefficient of repeatability was nearly twice as large for Rater 2 (2.96 mV) compared to Rater 1 (1.49 mV). However, reliability indices for MUNE were similar between the two raters (Table 2, CR was 55.5 and 56.2 for Rater 1 and 2, respectively), suggesting that the reliability of MScanFit MUNE was not strongly associated with rater reliability.

**Table 2.** Comparison of measures of reliability for the two primary outcome measures (CMAP and MUNE) for two-raters at the same site following the same protocol and using the same equipment. All measures use the M1 position for the reference electrode.

| Rater |  | Participant Sample |  |  |  | Visit 1 |  | Visit 2 |  | Relative and absolute reliability indices |  |  |  |  |  |  |  |
| --- | --- | --- | --- | --- | --- | --- | --- | --- | --- | --- | --- | --- | --- | --- | --- | --- | --- |
|  | Measure | N<br>(M, F) | Age<br>range<br>(years) | Mean<br>age | SD | Mean | SD | Mean | SD | ICC | Mean diff<br>(bias)<br>[95% CI] | SD<sub>diff</sub><br>between | p-value | 95% LOA<br>- lower | 95% LOA<br>- upper | Within<br>subject<br>variance | CR<br>(95%) |
| 1 | CMAP<br>(mV) | 17<br>(6, 11) | 18 - 75 | 46.4 | 18.7 | 6.91 | 1.30 | 6.92 | 1.31 | 0.83 | -0.01<br>[-0.36, 0.36] | 0.79 | 0.964 | -1.55 | 1.53 | 0.29 | 1.49 |
|  | MUNE |  |  |  |  | 147 | 30.6 | 152 | 19.7 | 0.38 | -4.4<br>[-8.48, 18.1] | 28.8 | 0.533 | -60.9 | 52.1 | 401.3 | 55.5 |
| 2 | CMAP<br>(mV) | 20<br>(14, 6) | 18 - 39 | 24.1 | 5.3 | 6.41 | 1.90 | 6.24 | 1.86 | 0.67 | -0.17<br>[-0.95, 0.39] | 1.54 | 0.640 | -2.85 | 3.19 | 1.14 | 2.96 |
|  | MUNE |  |  |  |  | 144 | 30.7 | 151 | 31.0 | 0.57 | -7.5<br>[-4.9, 19.8] | 28.4 | 0.255 | -63.2 | 48.2 | 411.9 | 56.2 |

N (M, F): number of participants, self-reported Male and Female.
ICC Intraclass correlation coefficient: two-way mixed effect model (absolute agreement definition).
Mean diff (bias): mean difference between visit 1 and 2, also known as bias in Bland-Altman plots, with 95% confidence interval.
SD<sub>diff</sub> between: standard deviation of the between subject difference in values at visit 1 and 2.
The reported p-value was generated by a two-sided permutation t-test (using 5000 bootstrap samples) comparing paired data from visit 1 and 2.
95% LOA - lower & upper: Bland-Altman 95% confidence intervals of the Limits Of Agreement, for lower and upper boundaries.
CR: Coefficient of Repeatability, 95% confidence interval, $=1.96 \cdot \sqrt{2 \cdot \text{within subject variance}}$

Pooling data from both cohorts, the average MUNE and coefficient of repeatability were estimated. Healthy participants exhibited an average MUNE of 148 (95% confidence interval 99 to 198, coefficient of variation 17.0%) and a coefficient of repeatability of 55 units. In an effort to assess the impact of MScanFit’s stochastic properties, the optimization process was repeated twelve times for each visit within the combined participant cohort. This iteration had no effect on the average MUNE or coefficient of repeatability (average 148 [95%CI 101 to 194], coefficient of variation 16.2%, CR 53).

### 4.0 Discussion

Our initial hypothesis, proposing that the alternative reference electrode position (A3, situated on the medial aspect of the patella) would represent the optimal choice for conducting MScanFit MUNE examinations of the tibialis anterior muscle, was not supported by the data. Instead, our findings suggest that the prescribed E2 electrode position outlined in the MScan Multicenter protocol (specifically, M1) emerged as the most suitable option, as supported by reliability indices. Additionally, it is noteworthy that the competence of the investigator performing the MScanFit MUNE examination does not appear to exert a substantial influence on the reliability of the final motor unit number estimate.

### 4.1 Electrically Silent E2 Reference Electrode Position

Our hypothesis regarding the optimal placement of the reference electrode (E2) on the medial aspect of the patella (A3 position) for conducting MScanFit MUNE investigations stemmed from the concept of electrical silence. Ideally, when recording the compound muscle action potential (CMAP) with a belly-tendon electrode configuration, the active electrode (E1) should be positioned at the motor point, while the reference electrode (E2) should be placed in a location that registers minimal, if any, evoked activity - essentially an “electrically silent” location. The A3 position demonstrated such electrical silence when subjected to supramaximal stimulation of the fibular (peroneal) nerve, as observed in prior studies (Escorcio-Bezerra et al., 2019; Nandedkar and Barkhaus, 2020). In contrast, the R1 position did not exhibit electrical silence, leading to reduced CMAP amplitudes both in Escorcio-Bezerra et al. (2019) and our present results (Figure 2).

However, intriguingly, CMAP amplitudes were comparable when the reference electrode was placed in either the A3 or M1 position (Figure 2). Given that the M1 position is near to several muscles innervated by the deep fibular nerve, including extensor digitorum longus, extensor hallucis longus, peroneus tertius, as well as the more distal intrinsic foot muscles such as extensor digitorum brevis and extensor hallucis brevis, we anticipated significant far-field potentials similar to those observed with the R1 position. We assessed electrical silence in all locations using prior methodology (Escorcio-Bezerra et al., 2019; Nandedkar and Barkhaus, 2020) and indeed detected measurable far-field potentials at the M1 position. However, practically speaking, these potentials were smaller than those recorded with the R1 position and did not consistently impact CMAP amplitudes measured from baseline to the early latency negative peak.

While it is important to acknowledge that our study did not systematically investigate CMAP latency and duration, post-hoc analysis validated our anecdotal observations that the CMAP latency and duration were shorter in the A3 position (Figure 1). The influence of the E2 position on CMAP latency and duration has been rigorously examined by Nandedkar and Barkhaus (2020, see their Figure 3). Their findings demonstrated that the proximal location for the E2 electrode (equivalent to our A3 position) resulted in shorter latency and duration for the CMAP compared to the distal position (which is about 5 cm distal to our M1 position). Again anecdotally, the shapes of the CMAPs during data collection were typically more *attractive* (to a neurophysiologist) when using the A3 position compared to the other positions. However, it’s important to acknowledge that the onset of the CMAP recorded with the A3 position consistently exhibited contamination by a significant stimulus artifact that was less prominent with the M1 position (Figure 1). While for motor nerve conduction studies, artifact reduction could be achieved by employing a shorter pulse width, minor adjustments of the bipolar stimulation probe, adjusting filter settings and, perhaps, adjusting the E0 electrode position, we opted to maintain these parameters in accordance with the MScan Multicenter protocol. Consequently, in the context of clinical investigations focusing on motor nerve conduction in the fibular nerve to the tibialis anterior muscle, the utilization of a proximal ‘A3’ position may still represent the most suitable choice.

### 4.2 Superior Reliability of the M1 Position

When deciding between the A3 and M1 positions for electrode placement, focusing solely on CMAP amplitude or MUNE values might not provide a definitive answer. However, an analysis of the relative and absolute reliability indices in Table 1 favors the M1 position due to its superior reliability metrics.

Intraclass correlation coefficients (ICC) reveal that CMAP and MUNE values exhibit moderate reliability with the M1 position, whereas with the A3 position, the reliability is moderate for CMAP but notably lower for MUNE (ICC = 0.25). Examining the coefficient of repeatability (CR) further underscores this difference. While CR values for CMAP are similar between the two positions, MUNE demonstrates smaller CR values when the M1 position is used (Table 1). This indicates that when using the proximal A3 position for the reference electrode, MScanFit MUNE assessments are associated with increased measurement error.

To explore this surprising result in more depth, we conducted the MScanFit optimization process on each study visit twelve times. This approach was based on sampling theory and the inherent random processes within MScanFit. By averaging the MUNE values from twelve runs, we aimed to identify the extent of variance in MUNE values attributed to the MScanFit process.

Interestingly, the impact on reliability metrics was minimal. For the M1 position, the ICC for MUNE improved slightly from 0.57 to 0.61, and the CR decreased marginally from 56.2 to 51.5 (Table 1).

This outcome aligns with the initial description of MScanFit by Bostock (2016). The optimization process has limited effect when the number of motor units is 80 or more, as demonstrated in simulated data (Bostock, 2016, Figure 5). Consequently, when dealing with healthy control participants with a substantial number of motor units, the MUNE values tend to converge, leading to the moderate reliability observed. This suggests that the primary source of variance affecting MScanFit’s reliability lies outside the optimization process for generating the “refined model.”

A serendipitous discovery emerged concerning the influence of the investigator’s competence on the reliability of the final motor unit number estimate. In cohort #1, the investigator exhibited a good intraclass correlation coefficient (ICC) of 0.83 and a coefficient of reliability (CR) of 1.49 mV for CMAP amplitude (Table 2). This implies that to consider a change in CMAP amplitude as a true change upon reexamination, it would need to exceed 1.49 mV; anything smaller could be attributed to measurement error. In cohort #2, the investigator achieved a moderate ICC of 0.67 and a CR of 2.96 mV. While examining the raw data from cohort #2, we observed six individuals with test-retest changes in CMAP amplitude that surpassed the limits of agreement (LOA, Table 2) established from data in cohort #1. Although no clear indications of errors warranted the exclusion of these participants, we retained all data, albeit with some suspicion regarding data quality. Nevertheless, this finding suggests that in the real-world context of multicenter studies and multiple investigators, MUNE reliability using MScanFit remains robust.

### 4.3 Limitations and Implications

One potential limitation of our data is the notable variability in CMAP amplitude observed in cohort #2. It could be postulated that more consistent primary data might have yielded different outcomes when using the proximal A3 position for the reference electrode. While this possibility exists, it’s important to note that the data from cohort #2 replicated the reduced CMAP amplitude observed when comparing the R1 to the A3 position, aligning with expected differences (Escorcio-Bezerra et al., 2019; Nandedkar and Barkhaus, 2020).

Now, considering the broader clinical implications, particularly in the context of monitoring individual patients with lower motor neuron degenerative disorders such as amyotrophic lateral sclerosis (ALS), several points merit discussion.

In our combined data from both cohorts of healthy controls, the average number of motor units estimated in the tibialis anterior muscle was approximately 148 (SD 25.2, 95% confidence interval 99 to 198). Suppose a patient presents in the early symptomatic phase of ALS with a MUNE value of 95 in the tibialis anterior. In this scenario, the MUNE value would be considered marginally low. However, when monitoring this individual over time, the MUNE would need to decline to a value of less than 40 to confidently attribute the change to a true drop in MUNE rather than variation due to measurement error. This is because the minimal detectable true change, represented by the coefficient of repeatability (CR), was calculated to be 55 units. This scenario raises concerns about the tool’s responsiveness, i.e., its ability to detect clinically relevant changes over time (Guyatt et al., 1987; Vaz et al., 2013).

It’s worth noting that this potential limitation is rooted in the assumption that the coefficient of repeatability, as determined in healthy controls, remains static as motor unit numbers decline. However, this is unlikely to hold true. Instead, the CR is expected to decrease as the mean number of motor units changes over time. To establish this dynamic value, further test-retest reliability studies are required, specifically involving individuals with deficits in motor unit number.

## Data Availability

All data produced are available online at Figshare.

https://doi.org/10.6084/m9.figshare.24112533.v1

